# Hybrid Value-Aware Transformer Architecture for Joint Learning from Longitudinal and Non-Longitudinal Clinical Data

**DOI:** 10.1101/2023.03.09.23287046

**Authors:** Yijun Shao, Yan Cheng, Stuart J. Nelson, Peter Kokkinos, Edward Y. Zamrini, Ali Ahmed, Qing Zeng-Treitler

## Abstract

Transformer is the latest deep neural network (DNN) architecture for sequence data learning that has revolutionized the field of natural language processing. This success has motivated researchers to explore its application in the healthcare domain. Despite the similarities between longitudinal clinical data and natural language data, clinical data presents unique complexities that make adapting Transformer to this domain challenging. To address this issue, we have designed a new Transformer-based DNN architecture, referred to as Hybrid Value-Aware Transformer (HVAT), which can jointly learn from longitudinal and non-longitudinal clinical data. HVAT is unique in the ability to learn from the numerical values associated with clinical codes/concepts such as labs, and also the use of a flexible longitudinal data representation called clinical tokens. We trained a prototype HVAT model on a case-control dataset, achieving high performance in predicting Alzheimer’s disease and related dementias as the patient outcome. The result demonstrates the potential of HVAT for broader clinical data learning tasks.

## 1. Introduction

Transformer is a deep neural network (DNN) architecture for learning from sequence data with natural language data as a primary example. Since its invention in 2017,^1^ Transformer has become the state-of-the-art approach in many natural language processing (NLP) tasks due to their excellent performance.^2-7^ The Transformer architecture is based on an attention mechanism, which allows the model to focus on different parts of the input sequence, thus capturing long-range dependencies and improving the quality of predictions.

One of the most successful applications of Transformer in NLP is the Bidirectional Encoder Representations from Transformers (BERT) model.^2^ BERT is a pre-trained language model that has achieved state-of-the-art performance on a wide range of NLP tasks, including text classification, question-answering, and named entity recognition. Another popular Transformer-based language model is the Generative Pre-trained Transformer (GPT),^3-5^ which has been used for various language generation tasks, such as machine translation and text summarization. The recently emerged chatbot known as ChatGPT, which has gone viral on the internet since its launch,^8^ was developed based on GPT to generate human-like responses to user input.^9^

The success of Transformer-based models in NLP has inspired researchers to explore their application in other domains such as healthcare. Longitudinal clinical data, which reside mostly in electronic health records (EHR), are also a type of sequence data, and in many ways similar to natural language data.^10,11^ In natural language, a sentence, for example, can be viewed as a sequence of words where the order of the words impacts its meaning. Thus, the positions of the words in a sentence are a critical component of the data. In clinical data, coded and/or textual data are recorded along with timestamps. Clinical concepts, which can be either defined using codes or extracted from the texts to represent diagnoses, medications, lab tests, vital signs, etc., play a similar role to the words in a sentence. The timestamps, which determine the temporal order of the clinical concepts, play a similar role to the word positions. Hence, it is a natural question whether the Transformer architecture can also be applied to clinical data and achieve excellent learning performance as well.

To answer this question, we must be aware that clinical data are much more complex than natural language data in several ways: 1) the time gaps between consecutive codes/concepts are irregular, while the position gaps between consecutive words are always one; 2) multiple codes/concepts may share the same time point, while no two words take the same position in a sentence; 3) a code/concept can have an associated numerical value (e.g., a lab test has a lab value), while no words have associated values; and 4) there are non-longitudinal data that are often needed for learning in addition to longitudinal data. These differences between clinical data and natural language data have made it a challenge to apply Transformer to clinical data for effective learning.

It is worth noting that there are Transformer-based clinical NLP models, which were obtained by fine-tuning the BERT model using clinical texts as specialized natural language data.^12,13^ These models are designed to deal only with unstructured text data, while we focus more on structured clinical data, including structured raw EHR data and data processed from both structured and/or unstructured raw data and presented in a structured form.

In recent years, several Transformer-based DNN architectures for structured EHR data learning were developed, which partially addressed the differences between clinical data and natural language data. Li et. al. proposed an architecture named BEHRT for pre-training using unlabeled data, which were further trained for disease prediction.^14^ Their model used only diagnoses and patient ages as input data. They represented patient data as a sequence of diagnostic codes, together with sequence orders of the visits and patient ages at the visits. Patient age was used both as a risk predictor and a source of temporal order information for the diagnoses. Their embedding layer used multiple types of embedding including concept embedding (for diagnoses), positional encoding (for visit sequence orders), age embedding, and segment embedding (for distinguishing adjacent visits). Rasmy et. al. similarly designed Med-BERT but for learning from a much larger vocabulary of diagnostic codes.^15^ In addition, they removed the use of ages, hence their data representation contained no temporal order information but only sequence order information. Pang et. al. introduced CEHR-BERT as an improvement on both BEHRT and Med-BERT.^16^ They expanded the vocabulary to include not only diagnoses but also medications, procedures, etc. and used both age embedding and time embedding to incorporate time information for each concept in the sequence. Kodialam et. al. proposed SARD architecture, which was also inspired by BEHRT.^17^ Their embedding is at the visit level, with each visit embedding being the sum of all embeddings of the concepts recorded during that visit. They used temporal encoding similar to the positional encoding in the original Transformer model to incorporate time information.

In this paper, we present a novel Transformer-based DNN architecture for joint learning from both longitudinal and nonlongitudinal clinical data. We referred to it as the Hybrid Value-Aware Transformer (HVAT). We also present a proof-of-concept experiment where a prototype HVAT model was developed using a dataset created in a prior study about Alzheimer’s Disease and Related Dementias (ADRD), to demonstrate the use and the capability of HVAT. Our design is different from the aforementioned architectures (e.g., BEHRT, etc.) in several ways. First, our architecture has a hybrid structure allowing for joint learning from both longitudinal and non-longitudinal data. Second, our architecture can learn from the clinical concepts/codes with numerical values from the longitudinal data. Third, we used a simpler but more flexible longitudinal data representation and embedding method.

## 2. Methods

### 2.1 HVAT Architecture

Based on the original Transformer architecture,^1^ we design the HVAT architecture for learning from both longitudinal and non-longitudinal clinical data. The HVAT architecture has two branches at the input end to receive longitudinal and non-longitudinal data, respectively (Figure 1). The main branch, taking longitudinal data as input, is called a Value-Aware Transformer (VAT). The second branch, taking non-longitudinal data as input, is a feed-forward neural network (FFNN) with a residual connection.^18^ The two branches join together at their last layers using a summation operation, followed by another FFNN with a residual connection, and lastly followed by the output layer, which is a single-node layer with the sigmoid function as the nonlinear activation function. The sigmoid function is defined as *σ*(*x*) = *e*^*x*^/(1+*e*^*x*^). The output is a single value *p* between 0 and 1. This type of output layer can be used for predicting binary outcomes coded by 0 and 1, respectively. The adverse outcome is usually coded by 1. The loss function is the binary cross-entropy function.

**Figure 1.**
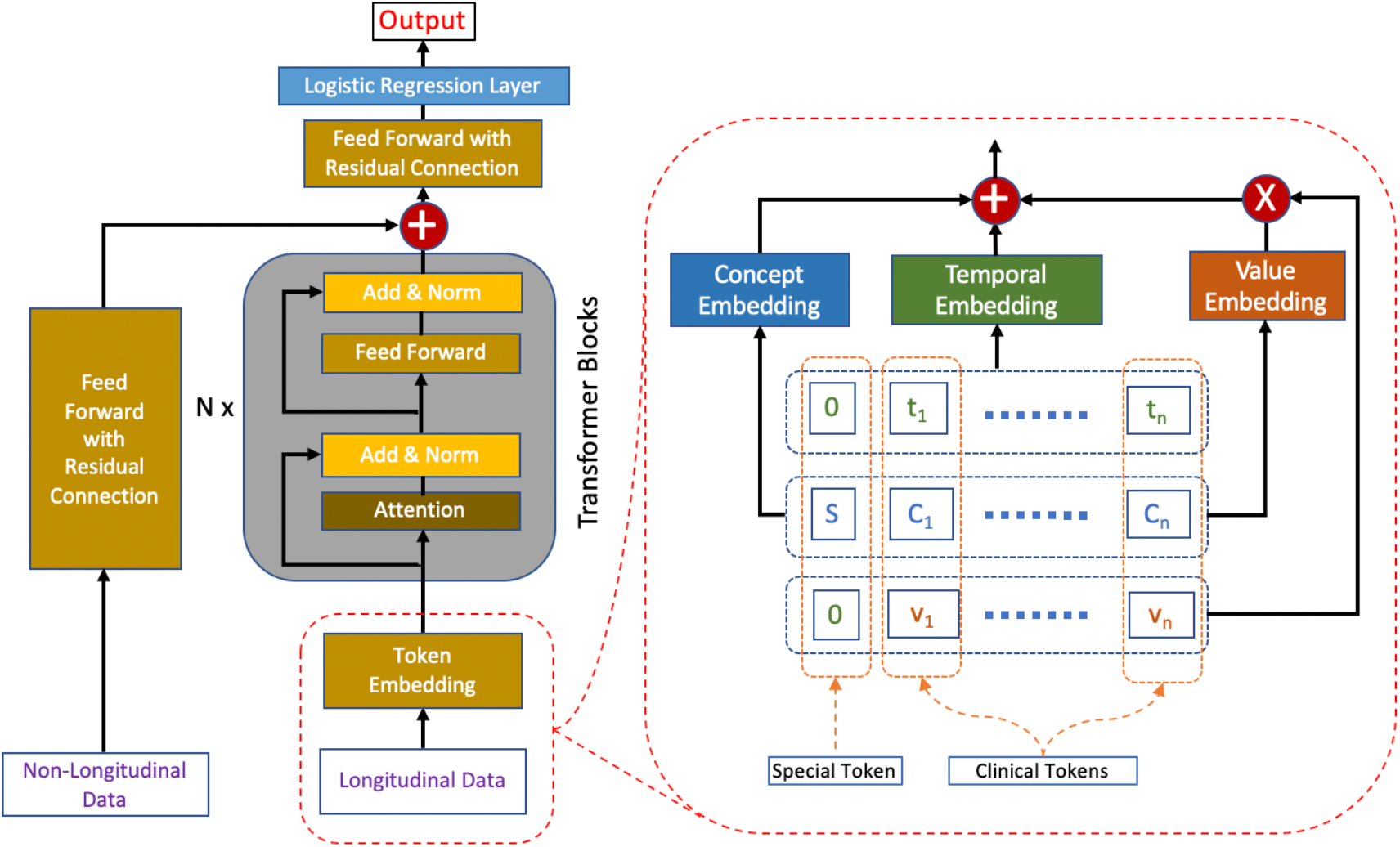
The HVAT architecture. The arrows indicate data flow directions. The left side (i.e., left to the words “Transformer Blocks”) is an overview of the HVAT architecture. The right side is a zoomed-in view of the part on the left side (enclosed by red dashed lines), which illustrates how the token embedding layer processes longitudinal data through three embeddings: concept embedding, temporal encoding, and value embedding.

### 2.2 Input Data Representation

Non-longitudinal clinical data are represented in the usual tabular format, i.e., one vector per patient, and the vectors are all of the same dimension. Such a representation is commonly used in traditional statistical modeling (e.g., logistic regression) and traditional machine learning (e.g., support vector machine).

For longitudinal clinical data, the representation is inspired by how the natural language data is represented and used by the original Transformer model. In natural language, a sentence is represented as a sequence of word tokens, and each word token is simply a word paired with its position in the sentence. Therefore, the repeated words in a sentence are considered different tokens. For example, in the sentence “to go or not to go”, there are 4 words (i.e., “to”, “go”, “or”, “not”) but 6 tokens:

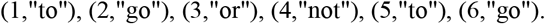

In general, word tokens generated from a sentence are written in the form (*i, w*), where *i* is the position of the word *w* in the sentence. Longitudinal clinical data are represented in a similar format through the following steps.

First, for each patient, we specify a time window for the patient history. We refer to the endpoint of the time window as the *index time/date*. The time window is divided into smaller equal-length intervals, and the intervals are indexed by natural numbers 1, 2, 3, …, from the latest to the earliest. These natural numbers are called *temporal indices*. The time window can vary from patient to patient, but the length of the small intervals is always the same for all patients. This step essentially normalizes the time information of the longitudinal data for all patients. How to choose the length of the intervals should depend on the specific tasks. For example, if the task is to predict outcomes for patients in an intensive care unit (ICU) stay, a good choice for the length may be at the scale from an hour to a day. If the task is to predict a disease that takes years to develop, then a good choice for the length may be at the scale from a month to a year.

Next, for each patient, the longitudinal data from the time window specified for the patient is represented as a sequence of clinical tokens, whose length may vary by patient. A *clinical token* is a triple (*t, C, v*), where *t* is a temporal index, *C* is a clinical concept, and *v* is either a numerical value associated with *C* or the default value zero. A *clinical concept* is defined as a clinically meaningful feature or variable identified or extracted from the clinical data. Examples of a clinical concept include: a disease, which may be represented by one diagnostic code or a group of related diagnostic codes, the prescription and use of a medication, a lab test, which may be represented by one lab code or a group of lab codes, a hospitalization, etc. Some clinical concepts have associated numerical values (e.g., lab tests have lab values) while others do not (e.g., diagnoses do not have associated values). Even if some clinical concepts have associated values in records, they can still be used as concepts without values. If *C* is a clinical concept with associated values, then *v* in the triple (*t, C, v*) is the associated value of *C* at temporal index *t*. If *C* is a clinical concept without values, then *v* takes the fixed value 0. If a concept *C* occurs multiple times over the same time interval indexed by *t*, only one token for this concept is generated for temporal index *t*, and if the multiple occurrences of this concept have multiple associated values, then the associated value *v* in the clinical token (*t, C, v*) is the value aggregated from the multiple values. The aggregation method depends on the particular concept and application. Below we give an example of representing longitudinal clinical data as clinical tokens.

Example: Suppose the time window for a patient is from 1/1/2012 to 12/31/2012, hence the index date is 12/31/2012. Within the time window, the patient had two diagnoses, Diabetes and Hypertension, on 1/19/2012; two lab tests, Calcium and Glucose, on 4/15/2012, with values 9.5 and 199, respectively; and three diagnoses, Diabetes, Hypertension and A-fib, on 10/8/2012. We choose the length of the time interval to be one month (or 30.5 days). Then the sequence of clinical tokens for this patient is:

(3, Diabetes, 0), (3, Hypertension, 0), (3, A-fib, 0), (9, Calcium, 9.5), (9, Glucose, 199), (12, Diabetes, 0), (12, Hypertension, 0).

Comparing the sequence of clinical tokens generated from the clinical data of a patient with the sequence of word tokens generated from a sentence in natural language, we see both similarities and differences (Table 1).

**Table 1.** A summarization of the similarities and differences between the representations of longitudinal clinical data and natural language data.

| Similarities | Differences |
| --- | --- |
| 1. The temporal index $t$ is analogous to the word position $i$ and the clinical concept $C$ is analogous to the word $w$.<br>2. The same clinical concept (resp. word) can occur at multiple but different temporal indices (resp. positions).<br>3. Every combination of temporal index (resp. position) and clinical concept (resp. word) can occur at most once in the sequence. | 1. Clinical concepts can have associated numerical values while words do not;<br>2. For a sequence of word tokens, the positions are consecutive integers with each integer occurring only once, while for a sequence of clinical tokens, the temporal indices from the sequence of clinical tokens are not necessarily consecutive integers: some integers can occur multiple times as temporal indices (though for different clinical concepts), and some integers may not occur at all. |

Therefore, the representation of longitudinal clinical data is similar to but also more complicated than the representation of natural language data.

For each patient, a special token is added to the sequence of clinical tokens generated from the patient’s data. The special token takes the form (0, *S*, 0), where *S* is an artificial concept defined to be different from all the clinical concepts. The output corresponding to the special token by the VAT branch will be a summarization of all the clinical tokens in the sequence, which is made possible by the attention layers in the Transformer blocks. Then this output, which is always a vector of a preset dimension regardless of the sequence length, and the output by the FFNN branch are summed together at the summation layer where the two branches join. Readers who are familiar with the BERT model will recognize that this special token plays the same role as the [CLS] token used in the BERT model.^2^

### 2.3 Token Embedding

When a clinical token (*t, C, v*) is fed into the VAT branch, it is first transformed by the token embedding layer into a vector of dimension *d*. The token embedding is defined based on three more basic embeddings: temporal embedding, concept embedding, and value embedding (Figure 1). Their specific definitions are given below:

1. The *temporal embedding*, which maps each temporal index *t* to a vector *TE*(*t*) of dimension *d*, is defined by

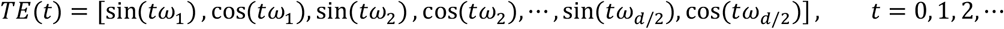

where *ω*_*k*_ = 10000^−2*k*/*d*^ for *k* = 1, …, *d*/2 (*d* is preset to be an even number). Note that the temporal embedding is not changed during the learning process.
2. The *concept embedding*, which is learned from the data, maps each concept *C* to a vector *CE*(*C*) of dimension *d*.
3. The *value embedding*, which is also learned from the data, maps each concept *C* to a second vector *VE*(*C*) of dimension *d*. The purpose of the value embedding is to provide a basis vector to incorporate the value associated with *C* into the token embedding.

The token embedding of the token (*t, C, v*) is defined by

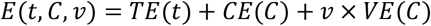

The token embeddings of all the tokens in a sequence are fed into the VAT branch simultaneously (Figure 1). In practice, *v* should be concept-wise normalized values rather than the raw values in the token embedding.

## 3. Experiment

### 3.1 Dataset and Cohort

The data source was the Veterans Affairs (VA) Corporate Data Warehouse, a nationwide EHR database for U.S. Veterans. We used the dataset created in a prior study which was focused on the association between physical fitness and ADRD risks.^19^ The study adopted the Exercise Treadmill Test (ETT) as the indicator for physical fitness, which was measured in Metabolic EquivalenTs (METs) (1 MET = 3.5 ml of oxygen utilized per kilogram of body weight). A natural language processing (NLP) model was developed^20^ and applied to all the medical notes in the database to extract MET values contained within those notes. For the initial cohort, we identified over 0.8 million patients who had at least one MET value between 2.0 and 23.9.

We employed a case-control design, where the cases were those who received an ADRD diagnosis, and the controls were those who did not. For the case group, we sampled from the initial cohort 50,000 patients who received an ADRD diagnosis on or before 12/31/2019. For the control group, we randomly sampled 50,000 Veterans from the initial cohort who were alive on 12/31/2019 and had never received any ADRD diagnosis by that date. The final cohort, having a total of 100,000 patients, was defined to be the combination of the case group and the control group. We defined the *endpoint* to be the date of the first ADRD diagnosis for each case and to be 12/31/2019 for each control. Then we defined the *index date* to be 3 months prior to the endpoint for everyone. The time window was defined to be from 1/1/2000 up to the index date. The time window was divided into intervals of one year long for the temporal index assignment.

To develop the HVAT model, we randomly split the cohort into three subsets: training (80%), validation (10%), and testing (10%), where each subset has an equal number of cases and controls.

### 3.2 Data Preparation

The outcomes were the case statuses, which were coded as: case = 1, control = 0. The predictors from non-longitudinal data included age (at index date), sex, race, and ethnicity. The predictors from longitudinal data included MET, Body-Mass Index (BMI), 15 diagnoses, 400 medications, and 400 note titles.

Age was used as a numerical variable. Sex was a binary variable coded as: female = 1, male = 0. Race was a multi-category variable with 4 categories: Black, White, Other, and Unknown. Taking White as the reference category, we converted race into 3 dummy variables named as: Race_Black (vs. White), Race_Other (vs. White), and Race_Unknown (vs. White). Ethnicity was a multi-category variable with 3 categories: Hispanic, Non-Hispanic, and Unknown. Taking Non-Hispanic as the reference category, we converted ethnicity to 2 dummy variables named as: Ethnicity_Hispanic (vs. Non-Hispanic), and Ethnicity_Unknown (vs. Non-Hispanic).

Among the features from longitudinal data, only MET and BMI were used as clinical concepts with values; all other features were used as clinical concepts without values. When there were multiple BMI values over the same time interval, we used the mean function for aggregation. When there were multiple MET values over the same time interval, we used the maximum function for aggregation. The 15 diagnoses were defined based on manually selected ICD-9-CM and ICD-10-CM codes. The 400 medications and 400 note titles were selected out of the total of thousands of medications and note titles using a feature selection method described below.

### 3.3 Feature Selection

Since both medications and note titles were used as clinical concepts without values, the information they carry is in their presence-absence status. For simplicity, we only considered the presence-absence status over the entire time window for each patient.

For each feature (a medication or a note title), we first calculated the 2×2 contingency table as follows:

|  | # in Cases | # in Controls |
| --- | --- | --- |
| Presence | $a$ | $b$ |
| Absence | $m - a$ | $n - b$ |

where *m* and *n* are the total numbers of cases and of controls, respectively. Features having distinct distributions in cases and controls are useful for prediction. For such a feature, the prevalence ratio (PR) defined as 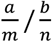 is expected to be distant from 1, or equivalently, the (natural) log of PR (LogPR) is expected to be distant from 0.

Next, we calculated the adjusted estimates of LogPR and its standard error SE, respectively, using the Walters formula:^21^

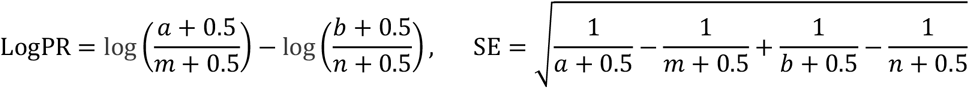

The equations were defined even when one of *a* and *b* was zero, thanks to the added small value 0.5. Then the confidence interval of LogPR at (1 − *α*)-level was

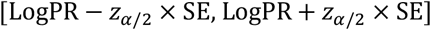

where *z*_*α*/2_ is the z-critical value corresponding to *α*/2.

We defined a feature ranking score using one of the two limits of the confidence interval: when LogPR ≥ 0, the score was the lower limit, and when LogPR < 0, the score was −1 times the upper limit. We found that this score could be expressed conveniently in one equation as

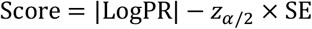

where | · | indicates the absolute value. Features with both a larger absolute value of LogPR and a smaller SE were ranked higher by this score. This score provides a balance between the two conditions, and the balance can be adjusted by changing *α*. A score less than 0 indicates that the LogPR is not significantly different from 0 at level 1 − *α*.

In this study, we estimated the feature ranking scores using the training set only, which meant *m* = *n* = 40,000. We chose *α* = 0.05, so *z*_*α*/2_ = 1.96. Then we selected the top 400 medications and the top 400 note titles ranked by the scores for model development. All of the selected features had a score greater than 0, which indicated that all of the selected features had a nonzero LogPR with statistical significance at *a* 95% confidence level.

### 3.4 Model Configuration

A HVAT model was developed using the final cohort to distinguish the cases from the controls. For the VAT branch, we used *N* = 2 Transformer blocks. We used multi-head attention with 2 heads within the Transformer blocks. For the token embedding, we set the dimension *d* = 32, which was also equal to the dimension of the output vector by the VAT branch. For all the dropout layers in the entire neural network, we set the dropout rate at 0.1. We used the rectified linear unit (ReLU) function^22^ as the nonlinear activation function in the entire neural network, except that for the output node, it was the sigmoid function.

### 3.5 Model Training and Evaluation

For model training, we set different learning rates for different parts of the HVAT model: 5×10^−4^ for the FFNN branch, 5×10^−5^ for the VAT branch, and 1×10^−4^ for the remaining part of the model. The weights in all the layers were initialized as small random numbers using K. He’s method.^23^ The weights were updated during training using mini-batch stochastic gradient descent with Nesterov momentum.^24^ The training set was divided into mini-batches consisting of varying numbers of patients. Patients within the same mini-batch had similar numbers of tokens in their respective sequences, and the total number of tokens in the mini-batch was equal or close to 10,000. Thus, the mini-batches with more patients all had shorter sequences and vice versa.

The primary metric of model performance was the Area Under the receiver operating characteristics Curve (AUC). The training process was stopped when the performance on the validation set plateaued, which was defined as no improvement in AUC over 10 consecutive epochs. The final model was set to be the one with the highest AUC before the plateau. Its performance on the testing set was reported as the model performance. The threshold on the output scores that maximized the accuracy on the training set was chosen to calculate the sensitivity, specificity, and accuracy on the testing set.

### 3.6 Ablation Study

In addition to the HVAT model, we also trained two other models to show how different the contributions of the longitudinal data and non-longitudinal data, respectively, are to the model performance. One model was the HVAT without the FFNN branch, which we named “Without FFNN”. This model only used longitudinal data as input. The other model was the HVAT without the VAT branch, which we named “Without VAT”. This model only used non-longitudinal data as input. Thus, we named the HVAT model “Full HVAT”.

### 3.7 Comparative Study

We also trained a linear support vector machine (SVM) model on the training set for comparison. The linear SVM model used the same predictors as the HVAT model, except that the presence-absence statuses and associated numerical values were all aggregated on the entire time window for all patients. Thus, the temporal information within the time window is lost.

## 4. Results

The performance on the testing set in AUC of the three models, i.e., Full HVAT, Without FFNN, and Without VAT, is shown in Figure 2.

**Figure 2.**
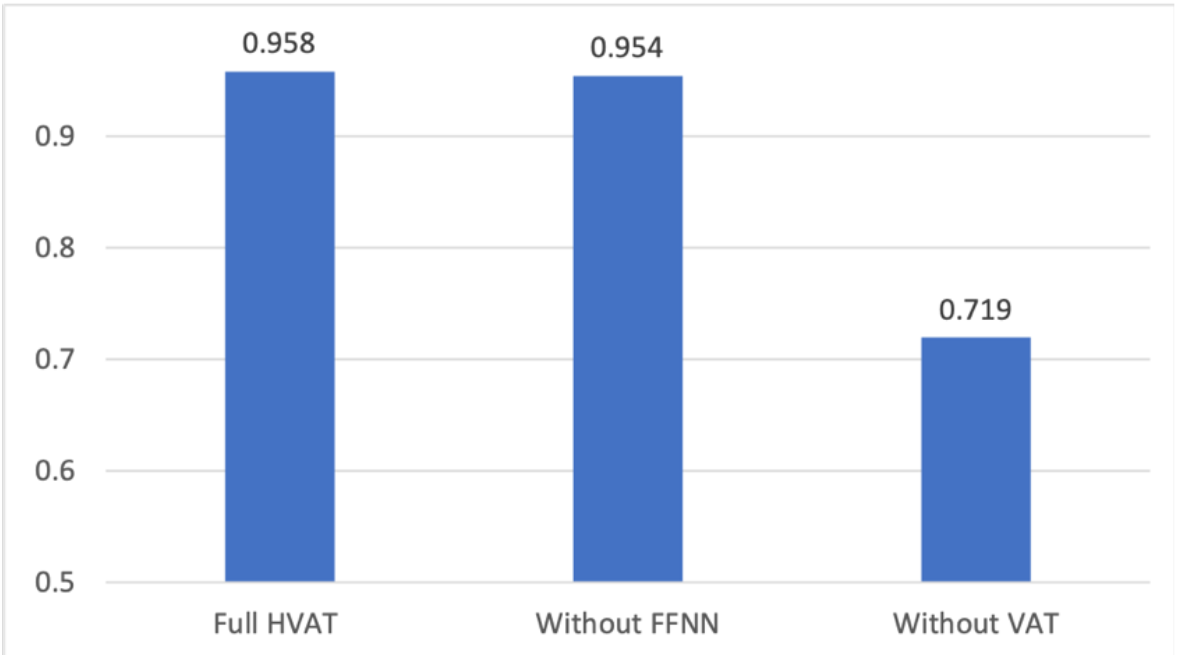
The performance in AUC of the three models: Full HVAT, Without VAT and Without FFNN.

The performance in AUC of the (Full) HVAT model and the SVM model on all three subsets is listed in Table 2.

**Table 2.** Comparison of the performance in AUC between SVM and HVAT.

| Model | Training Set | Validation Set | Testing Set |
| --- | --- | --- | --- |
| HVAT | 0.961 | 0.953 | 0.958 |
| SVM | 0.902 | 0.888 | 0.898 |

To report the performance of the HVAT model in other metrics, we first identified the threshold (= 0.6418) on the output scores that maximized the accuracy on the training set. Then we calculated the corresponding sensitivities, specificities, and accuracies on the three subsets using the threshold. The results are listed in Table 3.

**Table 3.** The performance of the full HVAT model.

| Metric | Training Set | Validation Set | Testing Set |
| --- | --- | --- | --- |
| Sensitivity | 0.863 | 0.852 | 0.853 |
| Specificity | 0.934 | 0.920 | 0.928 |
| Accuracy | 0.898 | 0.886 | 0.890 |

## 5. Discussion and Conclusion

In this study, we designed HVAT, a novel DNN architecture for joint learning from longitudinal and non-longitudinal clinical data. The HVAT was based on the original Transformer architecture designed for learning from natural language data, and our design leveraged the similarities while also addressing the differences between clinical data and natural language data. We further conducted a proof-of-concept experiment, in which a prototype HVAT model was developed to classify the patients in an ADRD cohort. The model achieved an excellent performance. The ablation study showed that the performance of the model with only the longitudinal data as input was very close to that of the HVAT model, while the model with only the non-longitudinal data was not so. This suggested that the high performance of the HVAT model was mainly due to the longitudinal data, which further showed that the model could learn from longitudinal data. The comparative study showed that the high performance of HVAT was not only due to the selection of good features but also the leveraging of the temporal information in the longitudinal data and its association with the outcome.

Compared to the other Transformer-based architectures for learning from clinical data, the HVAT is unique in that it can incorporate the numerical values associated with the clinical concepts as well as the non-longitudinal data with longitudinal data. The incorporation of numerical values in the architecture was made a key feature in our design because we realized that numerical values were common in clinical data and also crucial for outcome prediction. In our design, the values are incorporated into the token embedding as multipliers to the value embedding vectors, which is essentially equivalent to treating the clinical concepts with values as continuous variables. We admit that there is at least one other way to utilize the values: discretizing the range of values into multiple categories and then treating the concept with different categorized values as multiple different “concepts”. That way is similar to the common approach used in statistical modeling which treats a continuous variable as a multi-categorical variable by discretizing the continuous values. We did not adopt the latter approach because we realized that it has several disadvantages compared to our approach. First, the total number of tokens would be increased because of the multiple different “concepts” representing different values, which would further increase the computational burden including memory allocation and running time. Second, the multiple different concepts would be considered completely unrelated by the model, therefore, the association of one value with the outcome learned from data cannot inform the association of a nearby value with the outcome when the two values happen to fall into different categories. This would be a prominent problem for those categories of values having much fewer samples than others. Third, there is usually no standard way to discretize the continuous values, and many other considerations including both clinical and mathematical ones must be taken for doing that.

Another key component of our design of HVAT is the use of temporal indices for representing the occurrence time of the clinical concepts. The temporal indices essentially give an order for the time intervals within the time window of the patient history. There are two possible ways to order the time intervals: forward and backward. We chose the backward order because the time windows of the patients are supposed to be aligned along the end of the time window rather than the start, which is also why the end of the time window is defined as the index time/date. This way it makes the model easier to identify the relevant temporal patterns in the longitudinal data for predicting the outcomes.

The flexibility in customizing the length of the time intervals makes our architecture advantageous over the others. Actually, all the other architectures organize the longitudinal data at the visit level, which is equivalent to setting the time intervals to be one day long for HVAT. However, for some tasks such as the experiment we did in this study, where we used up to 20 years of history, lengthy sequences would be generated if the time intervals are set to be too short such as one day long. For example, assume an average patient has 10 visits per year and 10 concepts per visit, then over 20 years the patient would have 2000 tokens in the sequence. If we set the time intervals to be one year long, then the patient would only have 200 tokens in the sequence. Considering that the computational time of Transformer is O(N^2^) where N is the sequence length,^1^ our architecture would consume only 1% of the computational time of the others. Actually, it was exactly by setting the time intervals to be one year long that we were able to complete the model training and inference in a reasonable time on an ordinary desktop computer while an excellent performance was still achieved.

The ability to set the length for the time intervals also allows HVAT architecture to be useful in some situations where very short time intervals should be used. For example, in an ICU setting, the status of the patients may change from hour to hour, and to predict outcomes it may be best to set the time intervals to be one hour (or a few hours) long. It would be difficult for architectures designed for visit-level data structures to be applied to such situations.

Most of the other Transformer-based models were designed to work like the NLP model called BERT,^2^ whose main strength is its ability to pre-train the model without human labeling. The HVAT architecture also allows such pre-training although that was not demonstrated in our study. We believe that it is possible to design pre-training strategies similar to the “masked language model” and the “next sentence prediction” to pre-train a HVAT model on clinical data.

There are a few limitations in our study. First, we did not use certain important types of clinical concepts with values such as lab tests in the prototype model from the experiment. The lab values contain valuable information for outcome prediction. However, we find that the lab values are usually messier than other types of data such as diagnoses, and to use hundreds of lab tests as clinical concepts with values, it would take much effort and time to clean the lab data before they can be effectively used. Second, the prototype model is small in terms of the number of Transformer blocks, the dimension size of the embedding vectors, the number of attention heads, etc., compared to other Transformer-based models. This is mainly due to the limited computing resources available to us. However, the small model is sufficient for the purpose of prototyping.

For future work, we plan to develop explaining methods for the HVAT models which can reveal the relevant temporal patterns learned by the model and apply it to risk factor analysis for ADRD and other adverse outcomes. We are also contemplating using HVAT to train a GPT-like model as a foundational model which can generate simulated longitudinal clinical data.

## Data Availability

The raw patient-level data cannot be shared by policy, but aggregated data are available upon reasonable request to the authors

## Declarations

### Ethics Approval

This study has received an Institutional Review Board (IRB) exemption #1576748-1 from the Washington DC VA IRB.

### Funding

This study was funded by Grant NIH/NIA RF1AG069121 and Grant NIH/NIA 1R01AG073474-01A1.

## Notes

### Competing Interest Statement

The authors have declared no competing interest.

### Author Declarations

Ethics committee/IRB of Washington DC Veterans Affairs Medial Center gave ethical approval for this work

